# Investigating the feasibility and potential of combining industry AMR monitoring systems: a comparison with WHO GLASS

**DOI:** 10.1101/2024.03.27.24303768

**Authors:** Eve Rahbé, Aleksandra Kovacevic, Lulla Opatowski, Quentin J. Leclerc

**Affiliations:** Institut Pasteur, Université Paris Cité, Epidemiology and Modelling of Bacterial Escape to Antimicrobials (EMEA), 75015 Paris, France; INSERM, Université Paris-Saclay, Université de Versailles St-Quentin-en-Yvelines, Team Echappement aux Anti-infectieux et Pharmacoépidémiologie U1018, CESP, 78000 Versailles, France; Laboratoire Modélisation, Epidémiologie et Surveillance des Risques Sanitaires (MESuRS), Conservatoire National des Arts et Métiers, 75003 Paris, France

**Keywords:** antimicrobial resistance, surveillance, industry monitoring systems, GLASS

## Abstract

**Background:** Efforts to estimate the global burden of antimicrobial resistance (AMR) have highlighted gaps in existing surveillance systems. Data gathered from hospital networks globally by pharmaceutical industries to monitor antibiotic efficacy in different bacteria represent an additional source to track the temporal evolution of AMR. Here, we analysed available industry monitoring systems to assess to which extent combining them could help fill the gaps in our current understanding of AMR levels and trends.

**Methods:** We analysed six industry monitoring systems (ATLAS, GEARS, SIDERO-WT, KEYSTONE, DREAM, and SOAR) obtained from the Vivli platform and reviewed their respective isolates collection and analysis protocols. Using the R software, we designed a pipeline to harmonise and combine these into a single dataset. We assessed the reliability of resistance estimates from these sources by comparing the combined dataset to the publicly available subset of WHO GLASS for shared bacteria-antibiotic-country-year combinations.

**Results:** Combined, the industry monitoring systems cover 18 years (4 years for GLASS), 85 countries (71), 412 bacterial species (8), and 75 antibiotics (25). Although all industry systems followed a similar centralised testing approach, the criteria for isolate collection were unclear (patients selection, associated sampling periods…). For *E.coli*, *K. pneumoniae* and *S. aureus*, at least 65% of comparable resistance proportions were within 0.1 of the corresponding estimate in GLASS. We did not identify systemic bias towards resistance in industry systems compared to GLASS.

**Conclusions:** Combining industry monitoring systems can substantially strengthen our knowledge of global AMR burden across bacterial species and countries. High agreement values for available comparisons with GLASS suggest that data for other bacteria-antibiotic-country-year combinations only present in industry systems could complement GLASS, particularly for Priority Pathogens currently not covered. This valuable information on resistance levels could help clinicians and stakeholders prioritize testing and select appropriate antibiotics in settings with limited surveillance data.

**Plain language summary:** Antimicrobial resistance (AMR) is a growing problem worldwide, but we don’t always have enough information to fully understand its extent and how it’s changing over time. In this study, we looked at data collected by pharmaceutical companies from hospitals around the world to see how well antibiotics are working against different bacteria. We wanted to see if combining these data sources could help us fill in gaps in global AMR surveillance. We reviewed the methods of six different systems that collect this data and developed an approach to combine them. Then, we compared this combined data to publicly available GLASS data from the WHO to check if it was reliable. We found that the data from the pharmaceutical companies covered more years, countries, bacterial species, and antibiotics than GLASS. Even though the way the data was collected by the companies wasn’t always clear, we saw that the resistance estimates were similar to those from GLASS for some common bacteria like *E.coli*, *K. pneumoniae*, and *S. aureus*. Overall, combining data from these different sources could improve our understanding of AMR worldwide, especially in places where surveillance is currently limited, and for Priority Pathogens not covered by GLASS.

## Introduction

Implementing interventions to tackle the threat of antimicrobial resistance (AMR) first requires a good understanding of its global public health burden. Recent studies have highlighted multiple gaps in global AMR surveillance [1–3], which require new data sources to be addressed. Importantly, datasets must not only be summarised in reports, but also be publicly accessible and easily downloadable to facilitate further analyses by independent researchers.

Several initiatives have been developed to tackle AMR surveillance gaps. The most well-known include GLASS by the World Health Organisation [4] or EARS-Net by the European Centre for Disease Prevention and Control [5]. These initiatives provide standardised reporting guidelines to participating countries, and an infrastructure to collect and present aggregated AMR data. They currently only focus on a limited number of pathogens and antibiotics, but are informed by a substantial amount of isolates and are hence often referred to as reliable estimates of the prevalence of AMR. In parallel, several pharmaceutical companies conduct their own private global AMR reporting programs. These systems are designed to monitor drug efficacy in hospital settings by collecting a large number of isolates across countries and years and testing their susceptibility to a range of relevant antibiotics. Therefore, there may be an important role for industry programs to play in global AMR surveillance over time, as a complementary approach to public databases.

To the best of our knowledge, these different industry monitoring systems have been poorly explored and only separately, with no attempt to combine them yet. Combining these systems could broaden the range of pathogens, drugs, countries, and years covered, while also increasing the number of isolates used to inform AMR point prevalence estimates. This combination, however, requires a joint review of the surveillance methodologies of these different systems to clarify their similarities and differences. For example, clarifying how each system collects and conducts microbiological testing of isolates is crucial to determine the extent to which they can be combined and the potential biases they each have. Understanding the limits of different monitoring systems is an essential first step to appropriately utilise them.

Moreover, few studies have tried to compare AMR point prevalence estimates from different supranational surveillance systems [6–9]. It is important to know how AMR estimates from industry monitoring systems compare with publicly available initiatives. Agreement or differences between databases could reflect different sampling strategies, such as spatial coverage within a country or patient selection criteria. This information could be used to adapt sampling or coverage strategies, to provide better information to clinicians and stakeholders.

Here, we aim to clarify the value of industry AMR monitoring systems in tackling surveillance gaps worldwide. First, we evaluate the respective methodology of different systems including sampling process, patient selection, antibiotic susceptibility tests to determine if they could be combined and identify any challenges in this process. We then aim to assess the agreement of resistance proportions in these monitoring systems, individually or combined, compared to the publicly available subset of the WHO GLASS database.

## Methods

### Data acquisition

Data from Pfizer, GSK, Johnson & Johnson, Paratek, Venatorx, and Shionogi were obtained through https://amr.vivli.org. The GLASS dataset used in this study was obtained by merging two publicly available GLASS datasets obtained through different WHO sources. The first dataset was manually extracted from the WHO GLASS dashboard, introduced alongside the 2022 GLASS report (dataset available from https://github.com/qleclerc/GLASS2022). Importantly, this publicly available GLASS data does not include all the data used in the official WHO report [4], since it only presents data for countries which consistently reported isolates to GLASS for all years between 2017-2020. The data for some countries such as the United States is therefore not downloadable to the best of our knowledge. The second dataset is the complete GLASS dataset for 2019, included as supplementary electronic material alongside the 2021 GLASS report available from https://docs.google.com/spreadsheets/d/1Ej0a-av4V5uoFw19DfZoDvcLpdvHTscfXoqJgozGiwc/edit#gid=1592777314. We combined this dataset with the one extracted from the dashboard, which increased our coverage for 2019 from 46 countries and 2,547,754 isolates to 71 countries and 3,131,620 isolates. The combined GLASS dataset was then used for all the analyses presented in this article.

### Comparison of surveillance programs methodologies

The information about the methodology and spatio-temporal coverage of the available industry monitoring systems was acquired from the respective publications describing them [7,10–14]. Notably, we searched for information on criteria for collection of isolates, microbiological testing protocols, and reporting methods.

### Data reformatting and combination

To compare AMR estimates, we identified bacteria and antibiotics covered across multiple monitoring systems. We designed a flexible R script to convert minimum inhibitory concentrations (MIC) in the monitoring systems to resistant/susceptible labels using CLSI and EUCAST thresholds and aggregate AMR estimates across monitoring systems for any chosen combination of countries, years, bacteria, and antibiotics. All isolates were aggregated regardless of the sample source (blood, urine, stool etc…), as previous work has suggested resistance profiles are similar for commensal opportunistic pathogens across different sample sources [15].

To ensure comparability between GLASS and the industry monitoring systems, for bacterial species names, we assumed that AMR estimates in GLASS for “*Acinetobacter spp*” were representative of *Acinetobacter baumannii*. In industry monitoring systems, we assumed that a *S. aureus* isolate was considered to be methicillin-resistant (MRSA) if it was resistant to either methicillin, cefoxitin or oxacillin [5].

### Definition of a resistance proportion

Here, we defined AMR estimates using a “resistance proportion” metric, constructed as follows: the number of isolates labelled “resistant” over the total number of isolates tested (labelled “sensitive”, “intermediate” and “resistant”) for a given combination of bacterial species, antibiotic agent, country and year. This definition of resistance aligns with the updated EUCAST guidelines from 2021 [16].

### Comparison of resistance proportions

We adapted a previously published method [9] to calculate the “agreement” of resistance proportions among databases, using WHO GLASS as the reference [4]. The calculation involved determining the difference between resistance proportions in industry monitoring systems and those in GLASS. We derived both the average difference in resistance proportions and the proportion of comparisons with an absolute difference of less than 0.1. First, we compared each industry dataset to GLASS individually. Then, we combined all industry monitoring systems and assessed whether this improved the agreement.

Finally, we tested the relationship between the calculated resistance proportion differences and the number of isolates collected by the industry monitoring systems. Relationship was quantified using Spearman correlation coefficients.

### Code availability

The code developed for this project is available in a GitHub repository (https://github.com/qleclerc/AMR_data_prize). All analyses were conducted in R [17].

## Results

### Overview of industry monitoring systems methodology

All monitoring systems analysed here focus exclusively on invasive isolates [7,10–14]. However, since their primary purpose is to monitor drug efficacy, the focus of each system depends on the drug(s) monitored. ATLAS, GEARS, KEYSTONE and SIDERO-WT have a large coverage of antibiotics and bacterial species. On the other hand, DREAM exclusively aims to monitor bedaquiline efficacy and hence only focuses on multidrug-resistant *M. tuberculosis*. SOAR, in contrast, exclusively focuses on *S. pneumoniae* and *H. influenzae* (Table 1). Regardless of bacterial species, all systems except for DREAM gather isolates globally and send them to a single lab for MIC testing. ATLAS, GEARS, SIDERO-WT, and SOAR all use the services of the International Health Management Associates laboratory in the United States to conduct the MIC testing. This suggests that, in principle, the *in vitro* protocols are identical across these systems. On the other hand, DREAM sends MIC testing kits to participating labs and then relies on these labs to report their results. Lastly, while KEYSTONE explicitly distinguishes between isolates from hospital and community-acquired infections, and SOAR only represents community-acquired infections, other systems do not make this distinction. This lack of distinction may be problematic for pathogens that are known to display different resistance profiles depending on the infection setting [18,19].

**Table 1:**
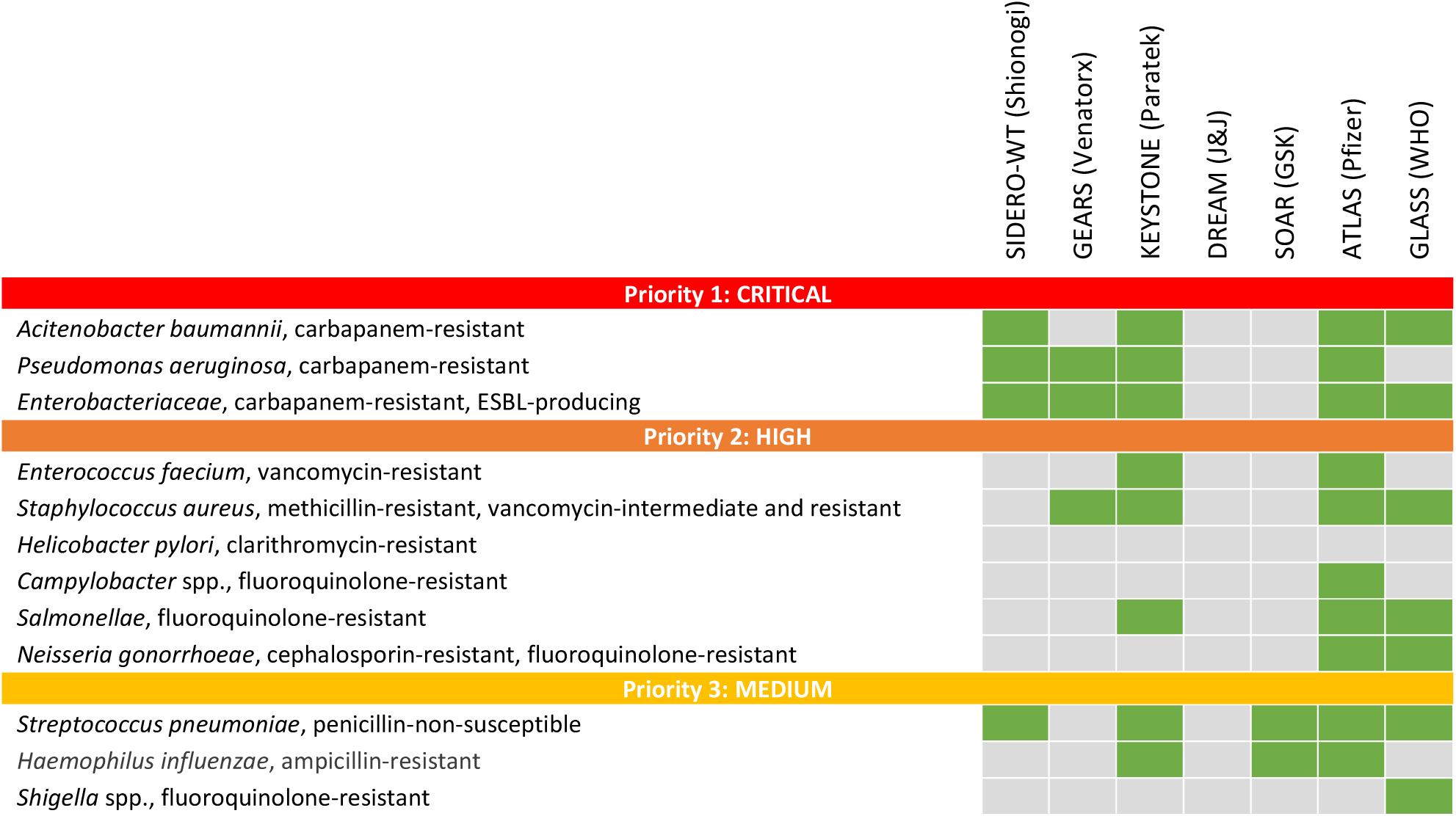
Bacteria-antibiotic availability across industry monitoring systems compared to WHO Priority Pathogens list (last updated in 2017). Green indicates presence and grey absence of the pathogen is in the corresponding monitoring system.

The major limitation common to all systems was a lack of clarity surrounding the selection of isolates for testing. In the SIDERO-WT program, isolates are randomly collected independently of resistance profile, following predetermined quotas for the number of isolates from different bacterial species to be collected at each participating centre [10]. In other systems however, even though the methodology briefly describes eligibility criteria, it is not clear whether all eligible patients are systematically enrolled or if there is a maximum number of patients. If there is a maximum, it’s unclear how these patients are chosen [7,11–14].

The coverage of each monitoring system is summarised in Table 2. In addition, we extracted the distribution of age groups covered in each dataset (Supplementary Figure 1). Although we were not able to compare with GLASS which does not include age, all industry monitoring systems have a similar distribution with isolates collected from individuals aged mostly between 19 and 84 years old. The exception is SIDERO, where 0-12 years old are better represented, at the expense of 65-84 years old.

**Table 2:** Individual dataset coverage. The numbers of countries, pathogens and antibiotics correspond to elements that appear at least once in the dataset, but not necessarily every year.

| Dataset | Years | Countries | Total isolates | Pathogens | Antibiotics |
| --- | --- | --- | --- | --- | --- |
| ATLAS (Pfizer) | 2004-2020 | 83 | 858,233 | 345 | 45 |
| GEARS (Venatorx) | 2018-2021 | 59 | 24,782 | 39 | 13 |
| SIDERO-WT (Shionogi) | 2014-2019 | 51 | 47,615 | 93 | 14 |
| KEYSTONE (Paratek) | 2014-2020 | 27 | 83,209 | 162 | 29 |
| DREAM (J&J) | 2011-2019 | 11 | 5,928 | 1 | 12 |
| SOAR (GSK) | 2014-2016 | 9 | 2,413 | 2 | 13 |
| GLASS (WHO) | 2017-2020 | 71 | 11,855,726 | 8 | 25 |

### Global coverage analysis

While the GLASS dataset analysed here covers 4 years, 71 countries, 8 species and resistance to 25 antibiotic agents (Figure 1a), the consideration of industry monitoring systems substantially increases the global coverage, as they encompass more years (18), countries (85), species (412) and antibiotics (75) (Figure 1b). We note multiple lower-resource settings covered by industry monitoring systems that are not included in the publicly available GLASS data, such as in the Americas, Central Europe or East Asia, despite current surveillance gaps in Africa still remaining, echoing previous work on this topic [2]. However, there are approximately ten times fewer total isolates in industry monitoring systems compared to GLASS (Figure 1c).

**Figure 1:**
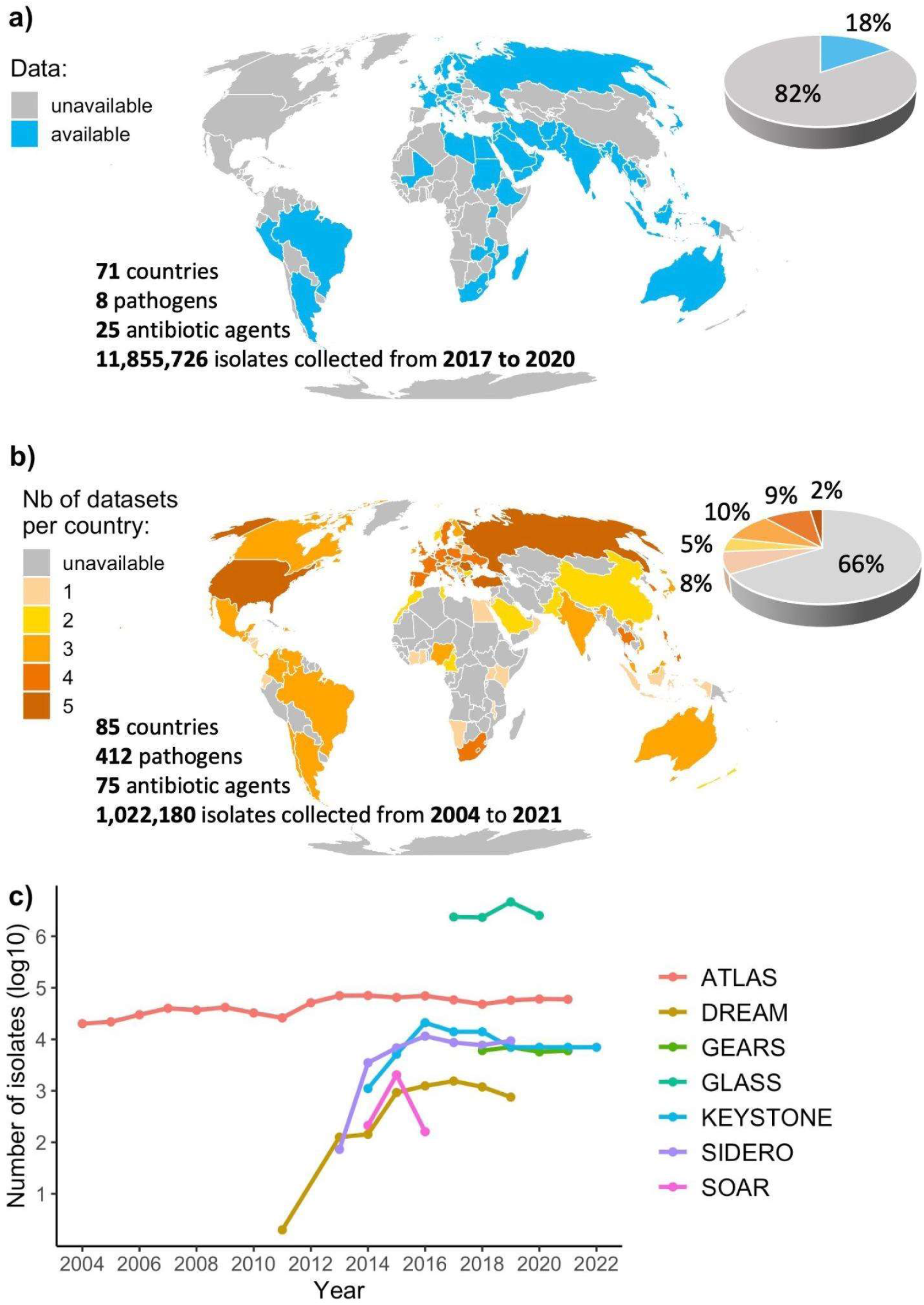
Global coverage of the surveillance monitoring systems. a) Coverage of the Global Antimicrobial Resistance and Use Surveillance System (GLASS). The coverage of GLASS presented here only includes data publicly available from the official WHO GLASS dashboard and supplementary data from the 2021 report, and therefore differs from the coverage presented in the latest 2022 report. **b) Combined coverage of six industry monitoring systems (ATLAS, DREAM, GEARS, KEYSTONE, SIDERO-WT and SOAR). c) Number of isolates per dataset per year .**

### Estimates of resistance across monitoring systems

For at least one common country and year, five bacterial species and 17 antibiotics were present in both GLASS and at least one industry monitoring system (31 unique bacteria-antibiotic combinations). *Salmonella* spp were also present in GLASS, ATLAS and KEYSTONE, but we excluded these bacteria from the analysis since there were less than 10 comparable isolates in the industry monitoring systems. *Shigella* spp were only present in GLASS but not in any industry dataset. Although some industry monitoring systems included *N. gonorrhoeae*, they did not cover the same years and countries as in GLASS, hence resistance proportions could not be compared.

We calculated resistance proportions by aggregating isolates by year, bacterial species and antibiotic to observe temporal trends. Within each bacterial species, antibiotics belonging to the same class had similar resistance proportions (Figure 2). The resistance proportions for *A. baumannii* are similar to those presented in a recent systematic review [20], except for tigecycline which is much higher here (between 0.5 and 0.75, compared to 0.15). The proportion of oxacillin-resistant *S. aureus* around 0.25 here (i.e. methicillin-resistant *S. aureus*) also falls within previously reported ranges [5,21]. Carbapenem resistance proportions for *E. coli* and *K. pneumonia* are similar to those reported in a recent systematic review (5% and 24%, respectively) [22]. Trends in resistance appear relatively stable, with the biggest changes seen between 2017 and 2018 (e.g. amikacin-resistant *A. baumannii* increase, trimethoprim/sulfamethoxazole-resistant *E. coli* and *K. pneumoniae* increase, ampicillin-resistant *E. coli* decrease, oxacillin-resistant *S. aureus* decrease).

**Figure 2:**
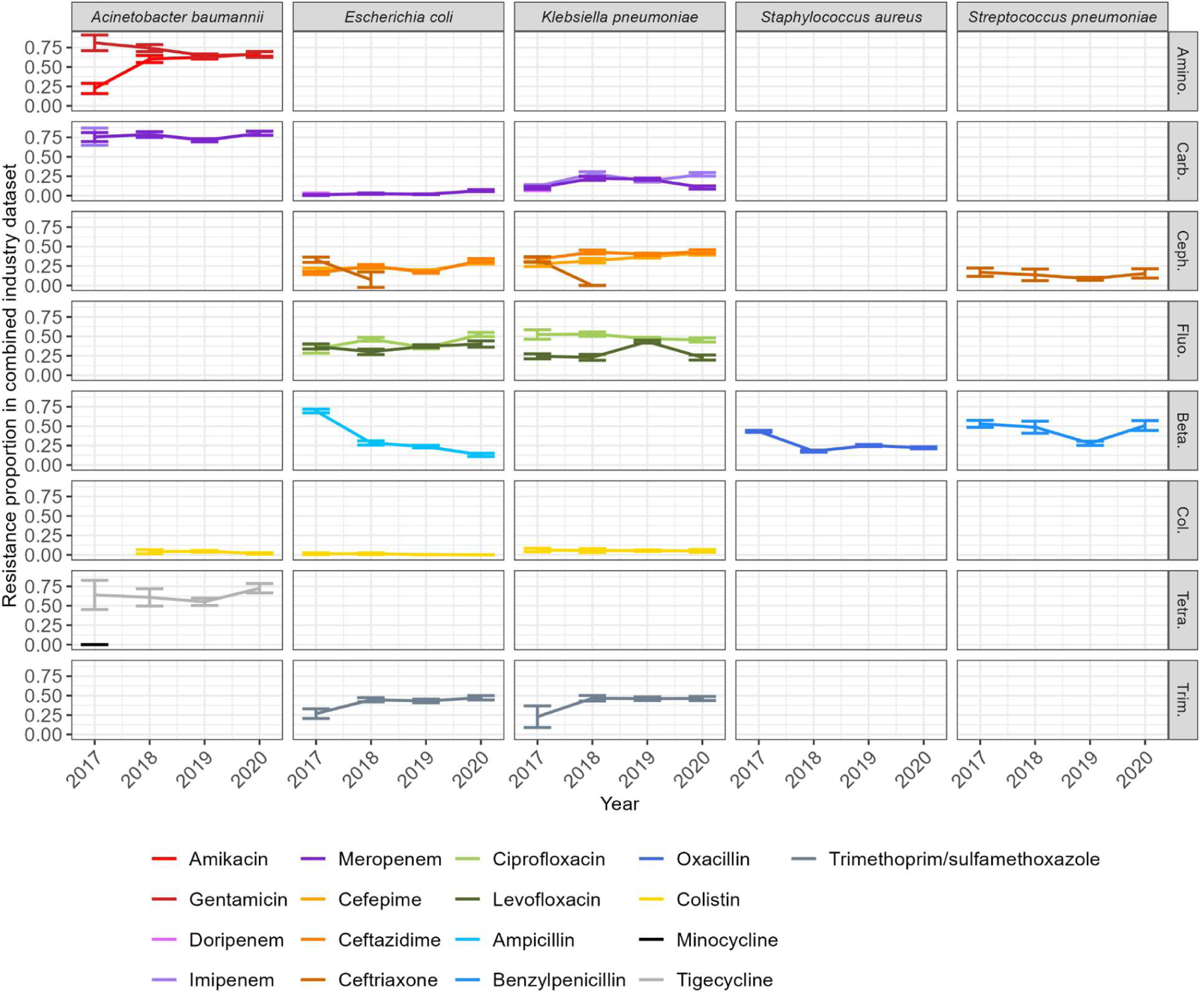
Resistance proportions by combinations of year-bacteria-antibiotics in the combined industry dataset. Here, isolates from different countries are aggregated to calculate resistance proportions. Confidence intervals indicate mean resistance +/- margin of error. Empty panels indicate absence of data for the corresponding bacteria-antibiotic combination. Antibiotics of the same colour but with a different shade belong to the same class. The classes represented are amino: aminoglycoside; carb: carbapenems; ceph: cephalosporins; fluo: fluoroquinolones; beta: beta-lactams; col: colistin; tetra: tetracycline; trim: trimethoprim/sulfamethoxazole.

The agreement between resistance proportions in GLASS and in the combined industry dataset varied between bacteria-antibiotic combinations (Figure 3). For *A. baumannii*, resistance was over-represented in the industry monitoring systems compared to GLASS, except for colistin. Interestingly, *A. baumannii* resistance proportions estimates greater than 0.6 for all antibiotics were mostly in agreement between the combined industry dataset and GLASS. Agreement was high for *E. coli*, *K. pneumoniae* and *S. aureus*, with 70%, 71% and 65% of all compared resistance proportions lying within +/-0.1 of each other, respectively. The exception was ampicillin for *E. coli*, for which resistance proportion estimates were under-represented in the industry monitoring systems compared to GLASS. Finally, resistance to both benzylpenicillin and ceftriaxone in *S. pneumoniae* were over-represented in industry monitoring systems compared to GLASS.

**Figure 3:**
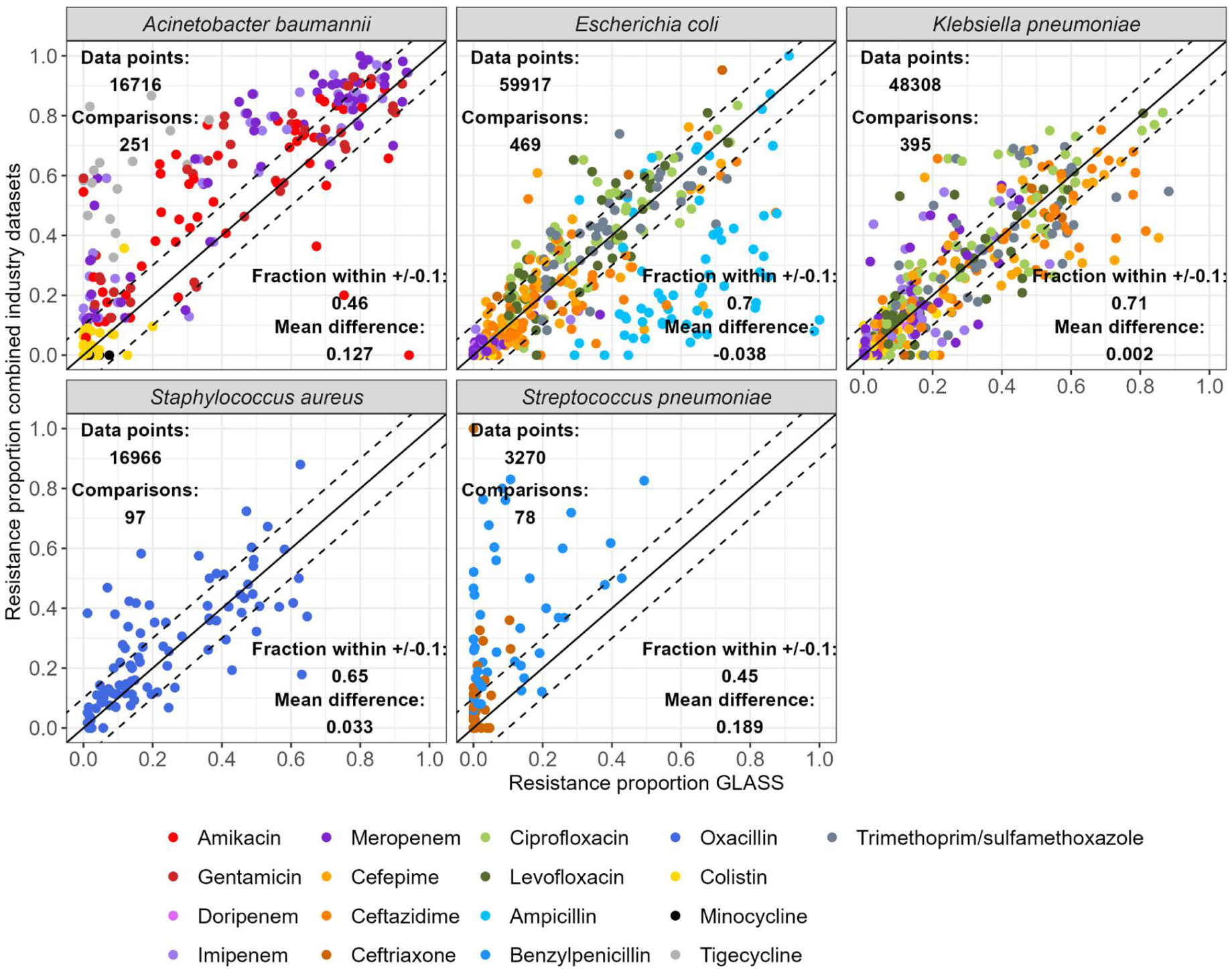
Comparison of resistance proportions by combinations of country-year-bacteria-antibiotics between the combined dataset and WHO GLASS. A “data point” is one resistance proportion result for one isolate (i.e. if a single isolate is tested for three different antibiotics, this adds up to three data points). A “comparison” is one combination of bacteria, antibiotic, country, and year found in both the combined dataset and GLASS (i.e. one point on the graph). Points on the solid line are comparisons where the proportion of resistant bacteria is identical in the industry and GLASS datasets. Points within the dashed lines are comparisons within +/-0.1 of each other. For individual industry monitoring system comparisons with GLASS, see Supplementary Figure 2.

**Figure 4:**
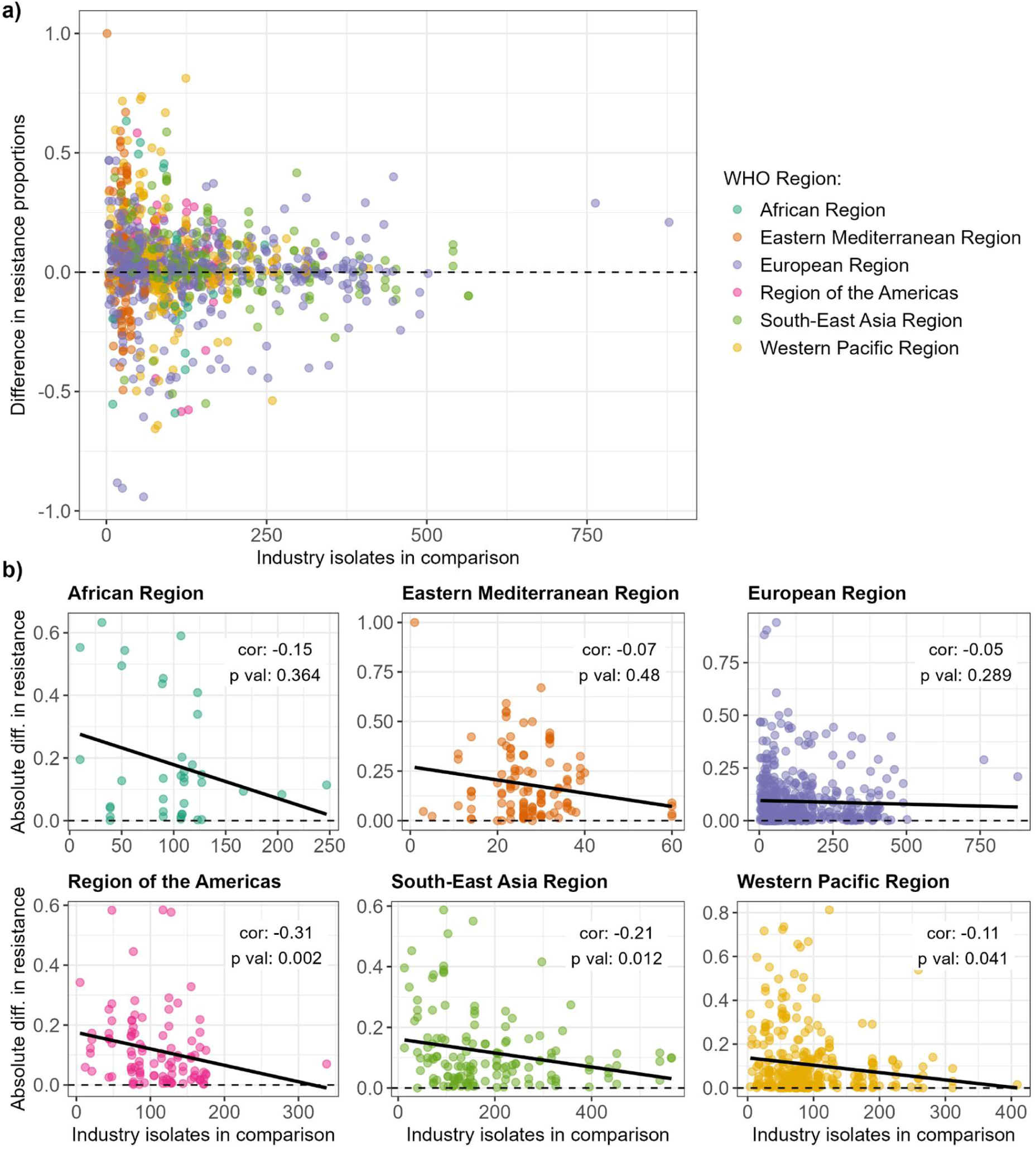
Relationship between resistance proportion difference between the combined industry dataset and WHO GLASS and number of isolates available from the industry dataset. a) Relationship for all WHO Regions. b) Relationships for each WHO Region separately. Spearman correlation coefficients and associated p-values are indicated on the graphs.

We also evaluated the agreement of individual monitoring systems with GLASS. ATLAS contained the most comparison points, but agreement of all monitoring systems compared to GLASS was good, with at least 45% of resistance proportions for any given combination of bacteria-monitoring systems within +/-0.1 of the equivalent estimate in GLASS (Supplementary Figure 2).

The difference between GLASS and combined industry dataset resistance proportions decreased as the number of isolates available in the industry dataset increased (Figure 4a). This observation applied to all WHO Regions, although the correlation was only statistically significant for America, South-East Asia and Western Pacific (Figure 4b; Spearman correlation, p value < 0.05 for significance).

Finally, we quantified the per-country agreement for each bacteria-antibiotic combination available for comparison (Supplementary Figures 3-7). The countries with the lowest agreement between the combined industry dataset estimates and GLASS estimates were not systematically those with a lower mean number of industry dataset isolates available to inform those estimates. Interestingly, some countries with the highest number of isolates had higher disagreements (e.g. *A. baumannii* in Malaysia in Supplementary Figure 3, *S. aureus* in India in Supplementary Figure 6, and *S. pneumoniae* in Japan in Supplementary Figure 7).

## Discussion

### Summary

Here, we demonstrate the potential value of merging industry monitoring systems originally aimed at monitoring antibiotic efficacy in different bacteria to increase the coverage of global AMR surveillance. The resistance estimates obtained from individual industry monitoring systems are similar to those from GLASS, where comparison is feasible and especially for *E. coli*, *K. pneumoniae* and *S. aureus*. Consequently, combining datasets in a relatively straightforward manner may not systematically result in a direct “increase” in agreement compared to the reference GLASS data, particularly in cases where the agreement between individual industry monitoring systems and GLASS is already high. The overall relatively good agreement suggests that resistance levels for many combinations of country-year-bacteria-antibiotic currently not covered in GLASS could be estimated from these industry monitoring systems. This is particularly important when attempting to improve our knowledge of AMR in lower-resource settings, and for Priority Pathogens that are not currently reported in GLASS (Table 1), such as *P. aeruginosa* (a critical priority pathogen included in four industry monitoring systems), *E. faecium* (considered high priority, included in two monitoring systems), and *H. influenzae* (listed as medium priority, found in three monitoring systems).

In agreement with previous findings [9], we observed that the greater the number of isolates tested to estimated resistance proportions by industry monitoring systems, the higher the agreement with GLASS. This suggests that resistance proportion differences between industry monitoring systems and GLASS may originate from limited data, rather than from a fundamental difference in the type of population from which isolates were sampled. However, this may not be the case for some specific countries, where we identified low agreement despite a relatively high number of isolates (Supplementary Figure 3-7). In such instances, this may indicate that healthcare institutions with substantially different characteristics are sampled in industry monitoring systems compared to GLASS.

We observed the highest disagreement for *S. pneumoniae*, where resistance to both benzylpenicillin and ceftriaxone was over-represented in industry monitoring systems compared to GLASS. Upon further inspection, we discovered that all data points of comparison for industry monitoring systems come solely from the ATLAS system (Supplementary Figure 2a). The ATLAS system lists sample sources but does not specify the type of pneumococcal disease, whether it is meningitis or non-meningitis. Knowing the type of infection is crucial for establishing resistance breakpoints for both benzylpenicillin and ceftriaxone, since non-meningitis infections have a high MIC breakpoint for both antibiotics (2 mg/L). In contrast, meningitis infections have lower MIC breakpoints of 0.06 mg/L and 0.5 mg/L for benzylpenicillin and ceftriaxone, respectively [23]. Therefore, assigning a resistance breakpoint may prove difficult in this case since it depends on the type of invasive pneumococcal disease, which may “overestimate” the resistance or report higher resistance than what we see in GLASS. The GLASS dataset also does not report the type of pneumococcal infection, but sensitive and resistant phenotypes have already been assigned.

### Limitations of monitoring systems

The main limitation in combining these industry monitoring systems was the challenge in identifying the criteria used for selecting the healthcare settings which provide the samples, as well as the criteria for selecting isolates for submission within these institutions. The selection process of sampled locations must be clarified to confirm the respective representativeness of monitoring systems within a country and to understand the differences in estimated resistance proportions across programs. For example, in cases where there is overlap between countries in different monitoring systems, it is not clear if each program collects isolates from different laboratories or medical institutions. In addition, understanding how isolates are sampled and chosen is essential to minimise the risk of bias towards either over- or under-representing resistant isolates. For example, if clinicians tend to select samples from patients for whom therapeutic failure was observed, this could lead to over-representation of AMR. Hence clarifying the isolate selection criteria will increase confidence in the value of AMR estimates.

Metadata on isolates should also be more systematically collected and harmonised. First, it would be helpful to distinguish between hospital- and community-acquired infections in those isolates, for greater insight into different AMR proportions in different settings. This distinction is generally made by identifying if the infection was reported within 48h of hospital admission (community-acquired) or later (hospital-acquired), hence information on patient hospitalisation date should be collected and compared to sample date. Second, sample source is a crucial information that is broadly collected but poorly standardised across monitoring systems. Within GLASS, sample sources are well categorised, clearly differentiating between urine, stool, genital or blood sources. However, these sources are not exhaustive, with respiratory isolates currently not included, for example. Although industry monitoring systems contain a much greater diversity of sample sources, the lack of harmonised labelling prevented us from including this element in our analysis. For example, the ATLAS system alone contains 97 unique labels for sample sources. This should be further explored since, in some cases such as for resistance rates in *S. pneumoniae*, analysing resistance proportions by sample source is necessary since MIC cut-off points vary accordingly.

The comparison with industry monitoring systems also highlighted several possible avenues for future development of GLASS. Firstly, available numbers of tested and resistant isolates are currently aggregated at the country-level. Increasing data availability by presenting phenotypic resistance at the isolate-level instead would enable us to better track the evolution of multi-drug resistances and allow finer comparison with industry monitoring systems. Indeed, while multi-drug resistance proportions could be calculated in the currently available GLASS version, a) only total numbers of isolates tested for each antibiotic are provided and b) not all isolates are tested for all antibiotics, there is a risk of bias due to double counting of aggregated isolates. Secondly, there is increasing interest in analysing minimum inhibitory concentration (MIC) data to observe finer trends in resistance evolution [9]. Susceptible/resistant labels currently reported in GLASS are clinically meaningful, but only relay binary information. Instead, asking countries to report MIC data to GLASS could allow finer analysis of resistance trends and earlier detection of abnormal variations. Thirdly, as explained in our Methods, here we only analysed the subset of the GLASS data that was publicly accessible from the GLASS website [4]. However, resistance proportions for several countries were not included in this publicly accessible GLASS subset, despite values for these same countries being presented in the official GLASS report. It is unclear to us why these countries are currently absent in the publicly available dataset, and may indicate differences in data sharing agreements.

### Next steps

To the best of our knowledge, this work is the first attempt to jointly investigate, compare, and combine all available industry AMR monitoring systems. We adapted the methodology from Catalán and colleagues [9], and applied it to multiple monitoring systems, using WHO GLASS as the reference dataset for comparison. This type of analysis is the essential first step for any work which aims to utilise these industry monitoring systems to their full potential. Without proper understanding of these methodological aspects and limits, the value of results cannot be trusted. Similarly, without the ability to combine these monitoring systems, we will miss opportunities to fill in gaps. Our approach can be repeated as new data are provided, to keep evaluating these monitoring systems going forward and iteratively suggest improvements. The code we developed to combine the monitoring systems is flexible and can be adjusted to select any combination of bacteria, antibiotics, and years of interest (https://github.com/qleclerc/AMR_data_prize).

In parallel to our analysis, WHO released in August 2023 an updated version of their manual guiding the implementation of GLASS [24]. Importantly, the bacterial species coverage will be extended to include two pathogens in the WHO Priority Pathogens list (*Pseudomonas aeruginosa* and *Haemophilus influenzae*), as well as *Neisseria meningitidis*, *Salmonella enterica* serovar Typhi, and *Salmonella enterica* serovar Paratyphi A. Four new types of sample sources will also be included (cerebrospinal fluid, respiratory samples, and rectal and pharyngeal swabs), which should facilitate future analyses of resistance stratified by sample source. It is unclear if these new guidelines will be implemented in time for the 2023 or even the 2024 report, but in any case, it will be interesting to revisit the comparisons we have made in our analysis using future versions of GLASS.

Overall, this work proposes a role for industry monitoring systems to fill-in known global surveillance gaps. We provide actionable points, suggestions, and comparison code for stakeholders to further improve these monitoring systems, with the aim to strengthen global health systems.

## Data Availability statement

The code and combined datasets used for this work are available in a GitHub repository (https://github.com/qleclerc/AMR_data_prize).

This project contains the following underlying data:

- **Publicly available GLASS data.** This dataset contains all GLASS data which, to our knowledge, can be publicly accessed from the 2022 report shinyapp and the 2021 report electronic supplementary material; note that this does not include all the data used in official GLASS reports. The data is available from https://github.com/qleclerc/GLASS2022.
- **Industry monitoring systems.** These are the six industry monitoring systems used in this analysis (ATLAS, GEARS, KEYSTONE, SIDERO-WT, SOAR). Access to these datasets can be requested from https://searchamr.vivli.org/.

Extended data are available online (https://doi.org/10.6084/m9.figshare.25408525). These contain single-dataset comparisons with GLASS, age distributions, and by-country agreement between the combined dataset and GLASS. These elements are presented as Supplementary Figures 1-7.

## Notes

### Competing Interest Statement

The authors have declared no competing interest.

### Funding Statement

This study did not receive any funding.

### Author Declarations

The study used ONLY openly available human data that can be obtained at: https://searchamr.vivli.org/ (industry monitoring systems) and https://github.com/qleclerc/GLASS2022 (GLASS data).

