## Extended Data for "Investigating the feasibility and potential of combining industry AMR monitoring systems: a comparison with WHO GLASS"

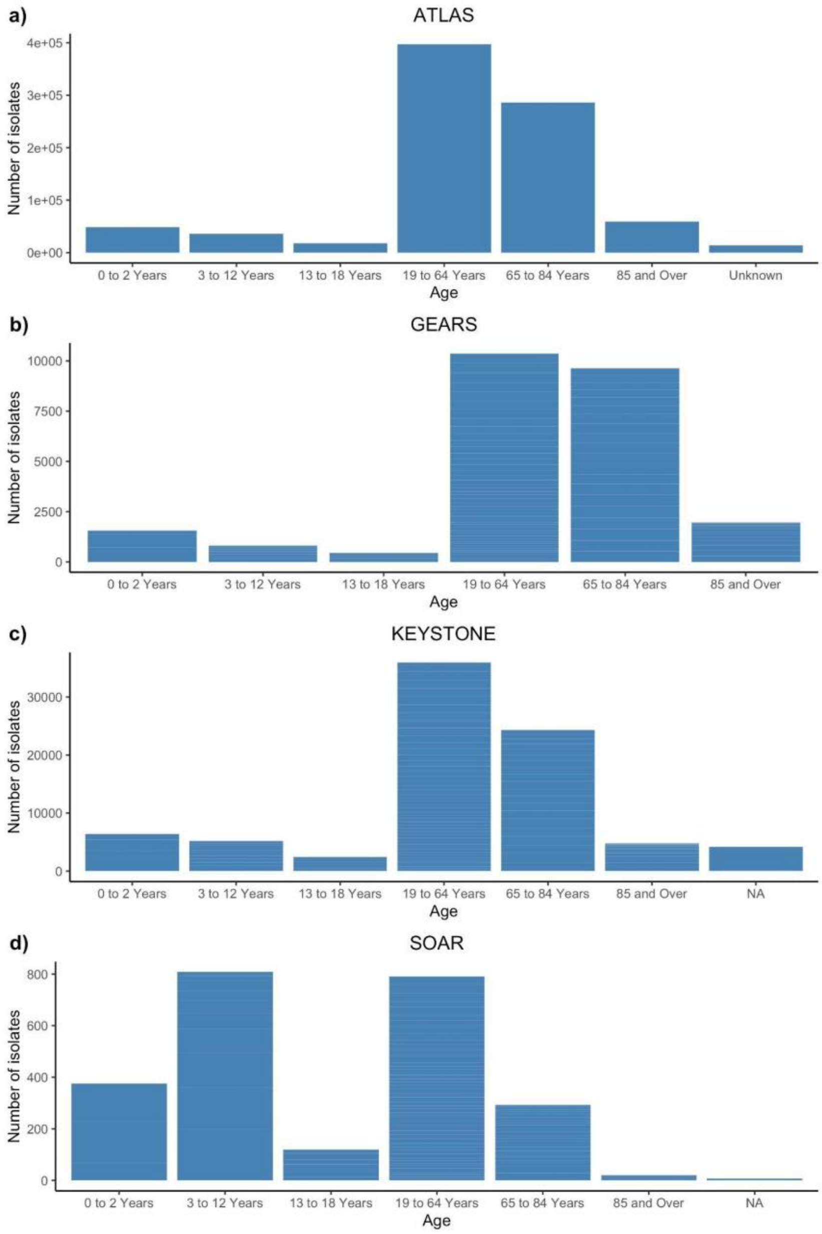


**Supplementary Figure 1: Age distribution of patients from which isolates were collected across industry monitoring systems. a) ATLAS, b) GEARS, c) KEYSTONE, d) SOAR**. SIDERO-WT, DREAM, and GLASS monitoring systems were not age stratified. SOAR has a greater proportion of isolates coming from children aged 0 to 12 years old compared to the other three monitoring systems.

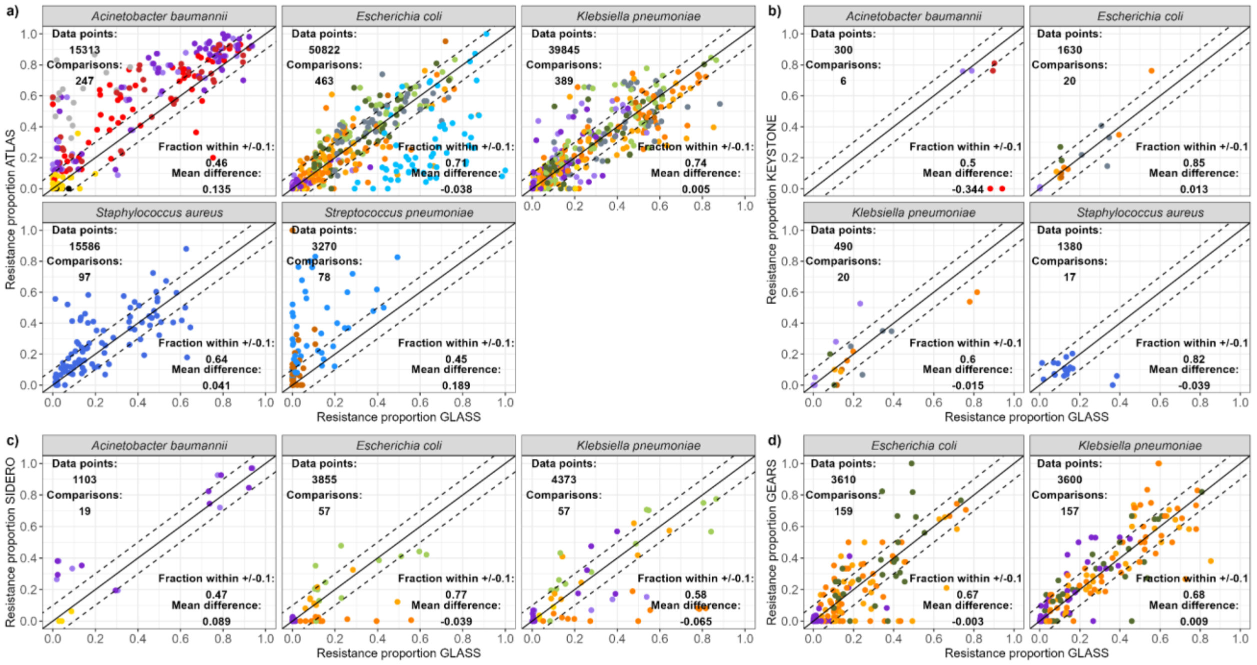


**Supplementary Figure 2: Comparison of resistance proportions by combinations of country-year-bacteria-antibiotics for each industry monitoring systems separately and WHO GLASS. a) ATLAS. b) KEYSTONE. c) SIDERO. d) GEARS.** A "data point" is one resistance proportion result for one isolate (i.e. if a single isolate is tested for three different antibiotics, this adds up to three data points). A "comparison" is one combination of bacteria, antibiotic, country and year found in both the industry monitoring system of interest and GLASS (i.e. one point on the graph). Points on the solid line are comparisons where the proportion of resistant bacteria is identical in the industry and GLASS datasets. Points within the dashed lines are comparisons within +/-0.1 of each other.

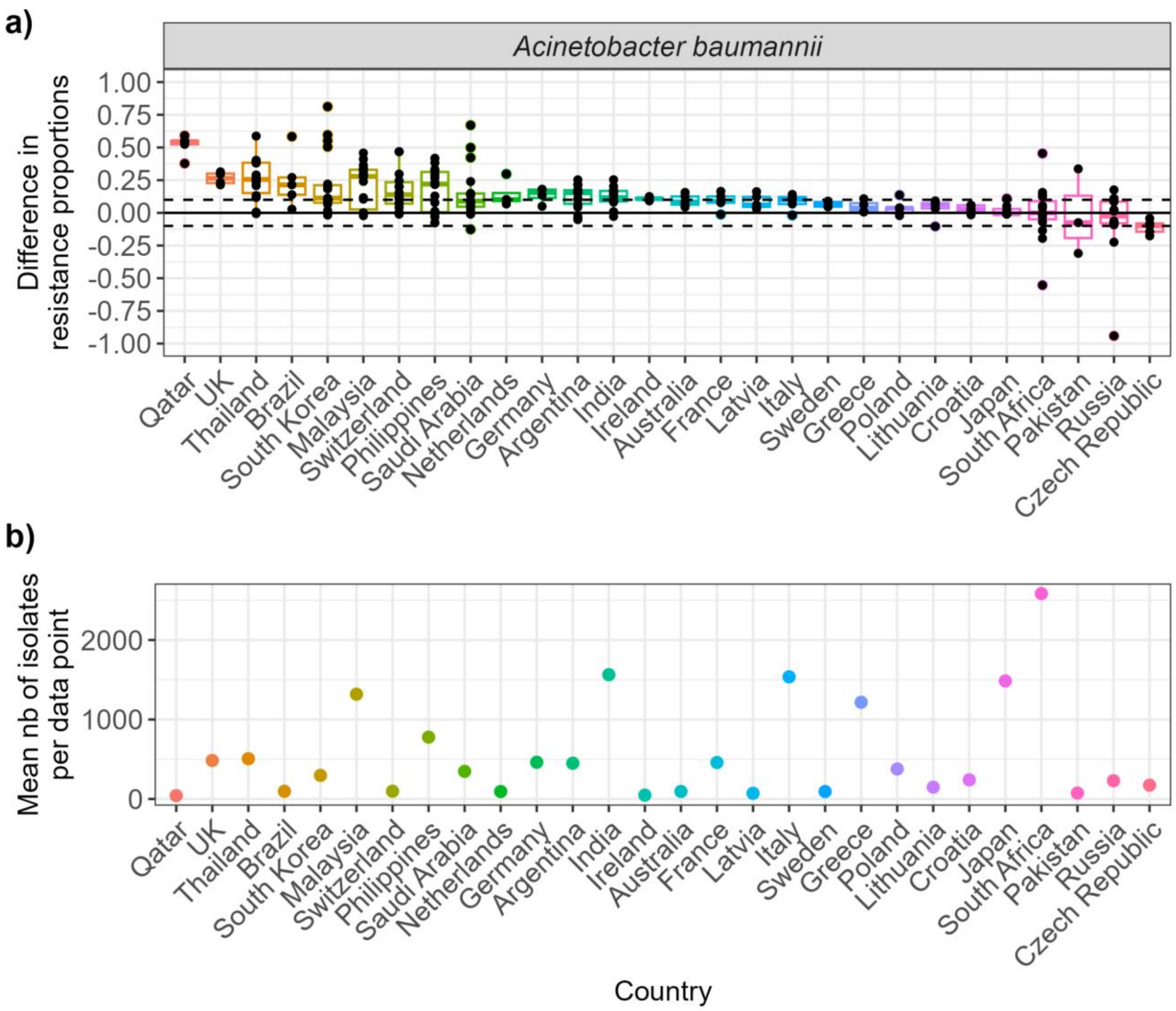


**Supplementary Figure 3: Comparing country-specific agreement on antibiotic-resistance values for *Acinetobacter baumannii*. a) Agreement between the combined industry dataset and GLASS.** Each point on the graph is one combination of bacteria, antibiotic, country and year found in both the combined dataset and GLASS. **b) Mean number of isolates per comparison point per country.** The values indicate the mean number of isolates available in the combined industry dataset to inform each comparison (i.e. each point on the graph in part a) of the figure).

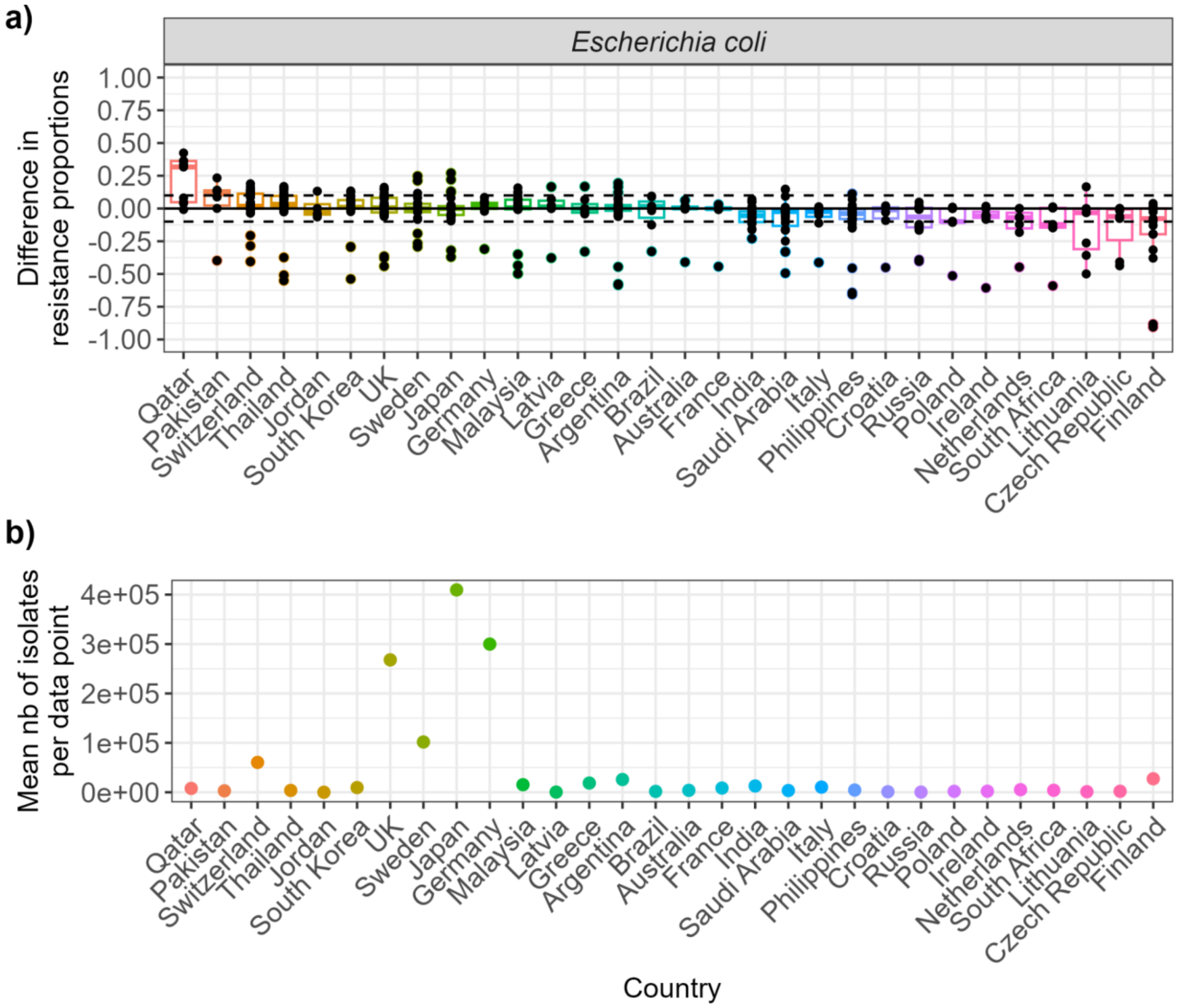


**Supplementary Figure 4: Comparing country-specific agreement on antibiotic-resistance values for *Escherichia coli*. a) Agreement between the combined industry dataset and GLASS.** Each point on the graph is one combination of bacteria, antibiotic, country and year found in both the combined dataset and GLASS. **b) Mean number of isolates per comparison point per country.** The values indicate the mean number of isolates available in the combined industry dataset to inform each comparison (i.e. each point on the graph in part a) of the figure).

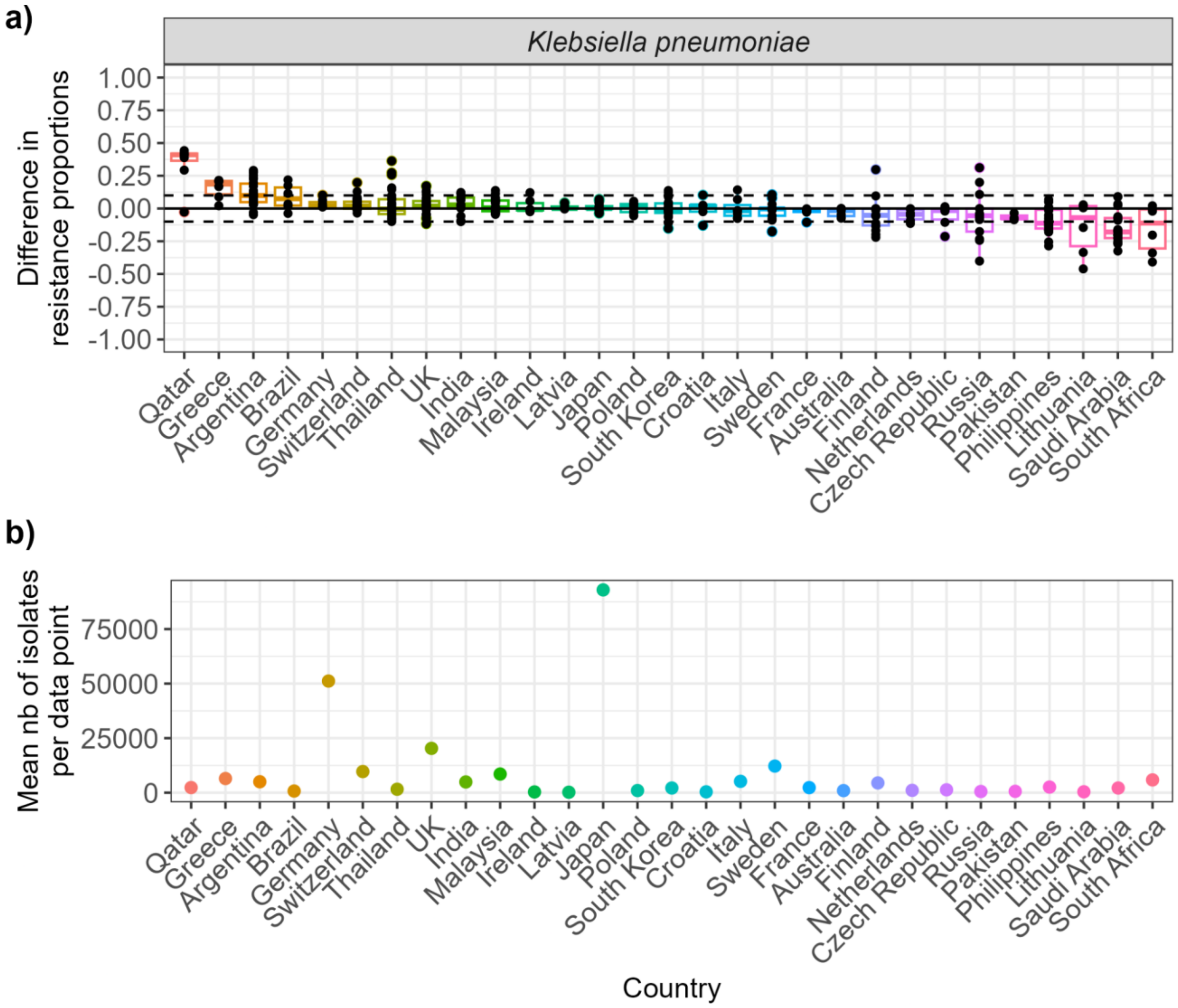


**Supplementary Figure 5: Comparing country-specific agreement on antibiotic-resistance values for *Klebsiella pneumoniae*. a) Agreement between the combined industry dataset and GLASS.** Each point on the graph is one combination of bacteria, antibiotic, country and year found in both the combined dataset and GLASS. **b) Mean number of isolates per comparison point per country.** The values indicate the mean number of isolates available in the combined industry dataset to inform each comparison (i.e. each point on the graph in part a) of the figure).

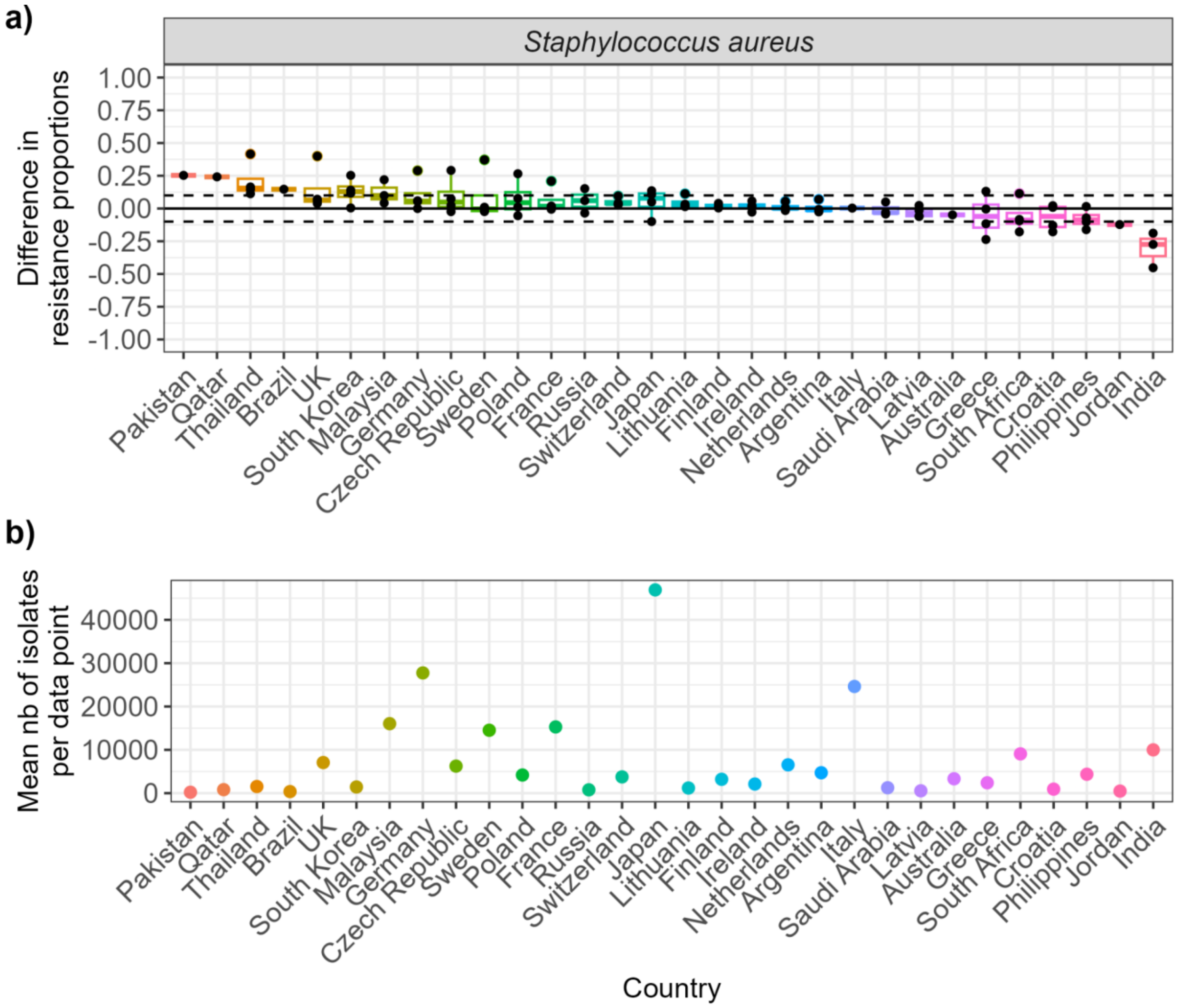


**Supplementary Figure 6: Comparing country-specific agreement on antibiotic-resistance values for *Staphylococcus aureus*. a) Agreement between the combined industry dataset and GLASS.** Each point on the graph is one combination of bacteria, antibiotic, country and year found in both the combined dataset and GLASS. **b) Mean number of isolates per comparison point per country.** The values indicate the mean number of isolates available in the combined industry dataset to inform each comparison (i.e. each point on the graph in part a) of the figure).

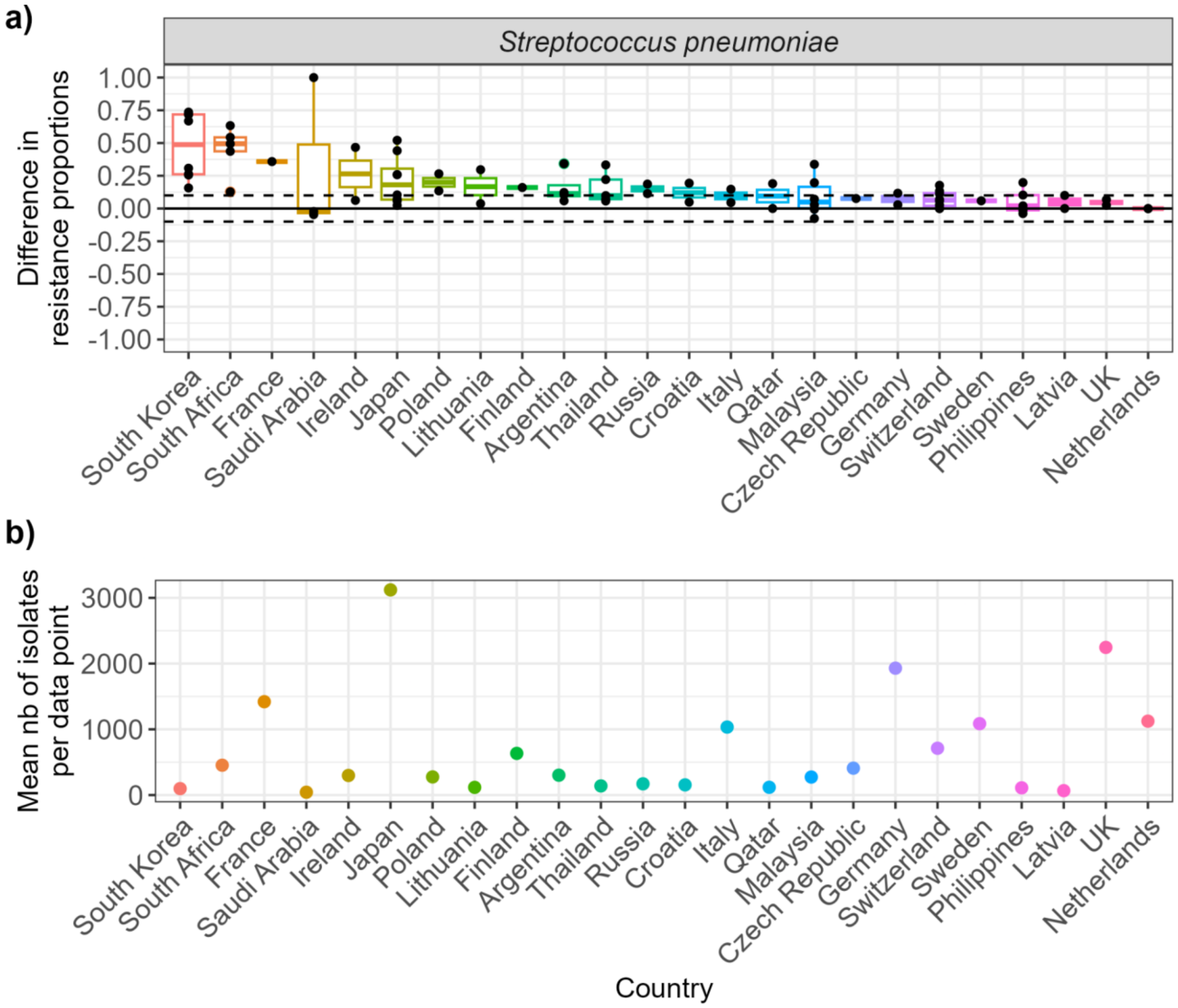


**Supplementary Figure 7: Comparing country-specific agreement on antibiotic-resistance values for *Streptococcus pneumoniae*. a) Agreement between the combined industry dataset and GLASS.** Each point on the graph is one combination of bacteria, antibiotic, country and year found in both the combined dataset and GLASS. **b) Mean number of isolates per comparison point per country.** The values indicate the mean number of isolates available in the combined industry dataset to inform each comparison (i.e. each point on the graph in part a) of the figure).
